# The Right Atrium Affects in silico Arrhythmia Vulnerability in Both Atria

**DOI:** 10.1101/2023.12.01.23299244

**Authors:** Patricia Martínez Díaz, Jorge Sánchez, Nikola André Fitzen, Ursula Ravens, Olaf Dössel, Axel Loewe

## Abstract

**Introduction:** The role of the right atrium (RA) in atrial fibrillation (AF) has long been overlooked. Computer models of the atria can aid in assessing how the RA influences arrhythmia vulnerability and in studying the role of RA drivers in the induction of AF, both aspects challenging to assess in living patients. It remains unclear if incorporating the RA influences the reentry inducibility of the model. As personalized ablation strategies rely on non-inducibility criteria, the adequacy of left atrium (LA)-only models for developing such ablation tools is uncertain.

**Aim:** To evaluate the effect of incorporating the RA in 3D patient-specific computer models on arrhythmia vulnerability.

**Methods:** Imaging data from 8 subjects were obtained to generate patient-specific computer models. We created 2 models for each subject: a monoatrial with only the LA and a biatrial with both the RA and LA. We considered 3 different states of substrate remodeling: healthy (H), mild (M), and severe (S). The Courtemanche et al. cellular model was modified from control conditions to a setup representing AF-induced remodeling with 0 %, 50 %, and 100 % changes for H, M, and S, respectively. Conduction velocity was set to 1.2, 1.0, and 0.8 m/s for each remodeling state. Fibrosis extent corresponded to Utah 2 (5-20 %) and Utah 4 (*>*35 %) stages for M and S, while the H state was modeled without fibrosis. Arrhythmia vulnerability was assessed by virtual S1S2 pacing from different points separated by 2cm using openCARP. A point was classified as inducing arrhythmia if reentry was maintained for at least 1 s. The vulnerability ratio was defined as the number of inducing points divided by the number of stimulation points. The mean tachycardia cycle length (TCL) was assessed at the stimulation site. We compared LA vulnerability ratios in monoatrial and biatrial models.

**Results:** Incorporating the RA increased the mean LA vulnerability ratio by 115.8 % (0.19 *±* 0.13 to 0.41 *±* 0.22, *p* = 0.033) in state M and 29.0 % in state S (0.31 *±* 0.14 to 0.40 *±* 0.15, *p* = 0.219). No arrhythmia was induced in the H models. RA inclusion increased the TCL of LA reentries by 5.5 % (186.9 *±* 13.3 ms to 197.2 *±* 18.3 ms, *p* = 0.006) in scenario M and decreased it by 7.2 % (224.3 *±* 27.6 ms to 208.2 *±* 34.8 *ms*, *p* = 0.010) in scenario S. RA inclusion increased LA inducibility revealing 5.5 *±* 3.0 new points per patient in the LA for the biatrial model, which did not induce reentry in the monoatrial model.

**Conclusions:** LA reentry vulnerability in a biatrial model is higher than in a monoatrial model. Incorporating the RA in patient-specific computational models unmasked potential inducing points in the LA. The RA had a substrate-dependent effect on reentry dynamics, altering the TCL of LA-induced reentries. Our results provide evidence for an important role of the RA in the maintenance and induction of arrhythmia in patient-specific computational models, thus suggesting the use of biatrial models.

## 1. Introduction

The role of the right atrium (RA) in atrial fibrillation (AF) has long been overlooked. Multiple studies have examined clinical conditions associated with AF, such as atrial enlargement, fibrosis extent, electrical remodeling, and wall thickening, but have been mainly concentrated on the left atrium (LA) [1, 2, 3, 4]. The focus on the LA in AF research can be attributed to two paradigms: First, it is now well established that the most frequent mechanism for AF onset is triggering activity from sources located in the pulmonary veins (PVs) of the LA [5]. The second paradigm refers to comorbidities associated with AF that primarily affect the left side of the heart, such as hypertension, valvular disease, and heart failure, which in combination increase mortality and reduce quality of life [6, 7]. Thus AF research continues to focus mostly on the LA, and as a consequence, the role of RA in AF is barely understood [8].

Computer models of the atria are becoming valuable tools for enhancing our understanding of intricate interactions during AF. These models provide a controlled and reproducible environment, enabling the study of specific research questions. For instance, these models can aid in assessing how the RA influences arrhythmia vulnerability and also in studying the role of RA drivers in the induction of AF, both aspects that would be challenging to assess in living patients until now. This work assesses the “Creative Concept” of incorporating the RA in computational arrhythmia studies.

Prior investigations tried to elucidate the role of the RA by proposing quantitative biatrial AF biomarkers. Using 3D transthoracic biatrial echocardiography, Soulat-Dufour et al. observed that AF remodeling is a biatrial process [9]. The same was observed invasively as a significant correlation in the remodeling state when comparing low-voltage areas in endocardial recordings from both chambers [10]. However, the findings reported by Chang et al. suggested that the LA had more areas of low voltage than the RA [11]. More recently, Hopman et al. showed that fibrosis burden in the RA is strongly correlated with the LA fibrosis extent based on late gadolinium enhancement (LGE) cardiac magnetic resonance imaging (MRI) [12]. Hasbe et al. described a case of paroxysmal AF initiated and maintained in the RA after adenosine infusion [13].

Another attempt to improve our understanding of the role of the RA in AF is to study sources beyond the PVs. Prior to the adoption of PV isolation as the standard treatment for AF, ablation of the RA was also investigated [14]. Several researchers have identified the presence of sources in the RA including the coronary sinus (CS), the superior vena cava (SVC), the right atrial appendage (RAA), the cavotricuspid isthmus (CTI), and the crista terminalis (CT) [15, 16, 17, 18]. To date, no approach was able to demonstrate consistent additional benefit when comparing the ablation of these regions with the ablation of PVs alone. The evidence for the role of the RA in AF remains inconclusive.

With the advent of personalized medicine, patient-specific computer models of the atria have already been used to identify ablation targets, tailor ablation strategies, and predict recurrence in AF patients [19, 20, 21, 22]. Nevertheless, those methodologies did not specifically focus on the role of the RA, with some excluding RA tissue and others neglecting the assessment of AF induction or maintenance from RA sources. In this study, we aim to evaluate the effect of incorporating the RA in 3D patient-specific computer models in the assessment of arrhythmia vulnerability based on 1398 virtual pacing sequences in 48 computational models.

## 2. Methods

A general overview of the study methodology is provided in Figure 1. Details on the 3 main steps are given in the following subsections.

**Figure 1:**
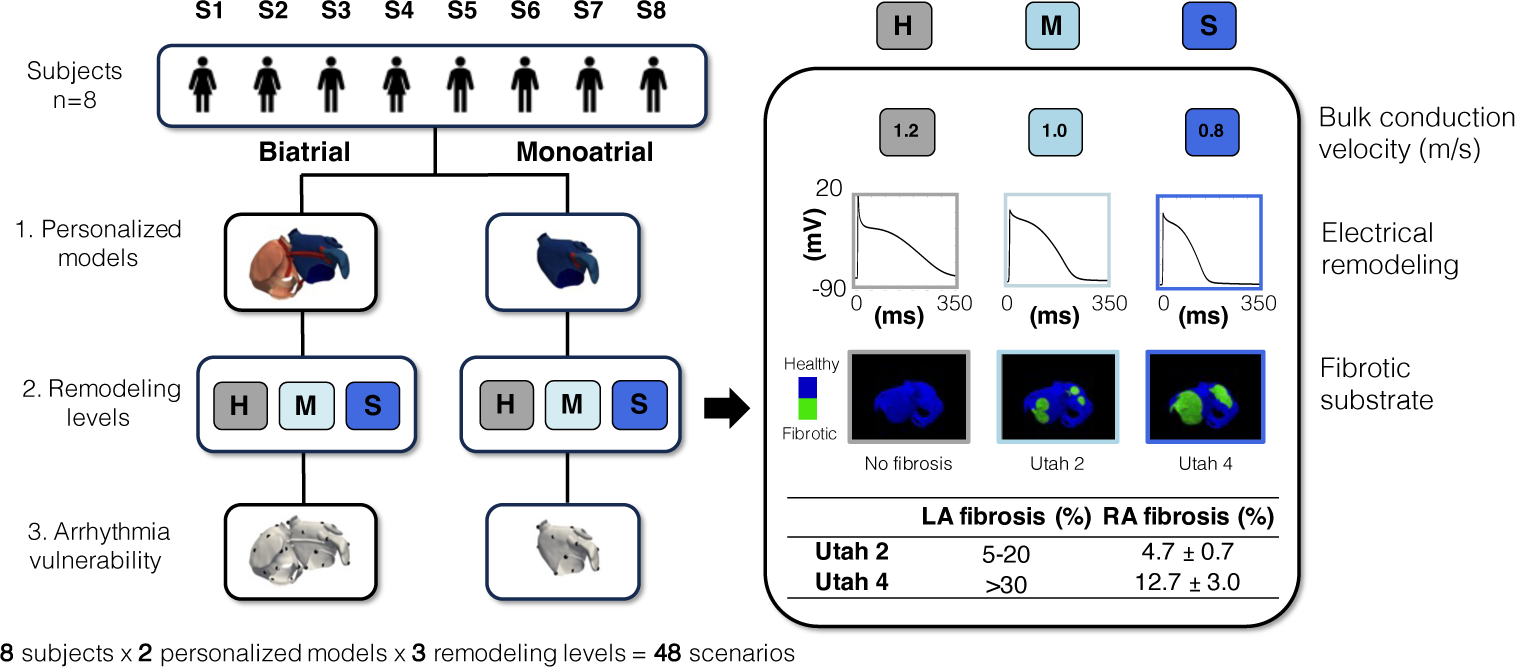
Study methodology. Left: generation of the virtual cohort considering (1) biatrial and monoatrial scenarios (2) at three different remodeling levels (3) for the assessment of arrhythmia vulnerability. Right: details of the fibrotic substrate modeling approach (H: healthy, M: mild, S: severe) considering changes in conduction velocity (CV), electrical remodeling, and fibrosis extent.

### 2.1. Patient-specific anatomical modeling

Imaging data from 8 subjects (S1-S8) were obtained as described in [23] and used to generate the biatrial personalized anatomical models. Subjects provided written informed consent and the study protocol was reviewed and approved by the ethical committee of Guy’s Hospital, London, UK, and University Hospital Heidelberg, Heidelberg, Germany. The cohort characteristics are described in Table S1 in the supplementary material. The personalized anatomical models were generated following the methodology described by Azzolin et al. [24]. Briefly, the veins and valves were manually removed from the endocardial surface, then the meshes were resampled to an average edge length of 400 µm. Subsequently, 4 inter-atrial connections (IAC) were automatically generated to link the RA and LA [25]. Next, a rule-based approach based on the solution of Laplacian equations was employed to define preferential myocyte orientation. Multiple anatomical regions such as the CT, pectinate muscles (PM), and Bachmann’s bundle (BB) were automatically annotated to consider electrophysiological heterogeneity [26]. Finally, to take into account wall thickness, bilayer meshes were generated by extruding the endocardium and connecting it to the epicardium as described in [27]. For each subject, we created two models: one consisting only of the LA, which we will refer to as *monoatrial*, and one model consisting of both the RA and LA, which we will refer to as *biatrial*.

Cellular electrophysiology of atrial myocytes was modeled using the mathematical model of Courtemanche-Ramirez-Nattel (CRN) [28]. To compute electrical propagation in the human atria we solved the monodomain equation using the electrophysiology simulator openCARP [29, 30]. A carputils bundle containing the openCARP experiment is publicly available [31].

### 2.2. Electrophysiological modeling

We defined 3 different levels of atrial fibrillation-induced remodeling, namely: healthy (H), mild (M), and severe (S) by reducing the conductance of a set of ionic channels as described in [32] with 0 %, 50 % and 100 % changes for H, M and S, respectively. The maximum scaling of the ionic conductances has an effect on the action potential in line with the changes observed in human atrial myocytes in patients suffering from persistent AF [33]. The model was parameterized to yield a conduction velocity (CV) along the myocyte preferential direction of 1.2, 1.0, and 0.8 m/s for each remodeling level, respectively. Intra- and extracellular conductivities were scaled x 3 for the BB and x 2 for the CT and PM. Anisotropic wave propagation was modeled using regional ratios as in [34].

### 2.3. Fibrotic substrate modeling

The fibrotic substrate was modeled following the methodology described by Nagel et al. [35]. Briefly, we manually placed 6 seeds on each biatrial model, 3 in the LA and 3 in the RA, which correspond to regions with most frequent enhancement (IIR*>*1.2) in LGE-MRI [36, 37, 38, 39]: the posterior LA wall close to the inferior PV, the inferior wall in proximity to the inferior PV, and the lateral wall. In the RA: close to the IVC junction, the septal wall, and in the venous component (see Figure S1 in the supplementary material). The Utah stage classification [40] was used to define the ranges of fibrosis in each remodeling level. The M and S fibrotic substrate levels corresponded to the Utah 2 and Utah 4 stages, respectively. The proportion of RA and LA fibrosis extent was based on the percentages reported by Akoum et al. [41]. The fibrotic regions were modeled with 30% of the elements non-conductive and the rest with TGF-,*8*1-induced electrical remodeling [20, 42, 43].

### 2.4. Arrhythmia vulnerability

Arrhythmia vulnerability was assessed by an S1-S2 pacing protocol [44] from a set of stimulation points with 2 cm inter-point distance on the atrial surface. Stimulation points and earliest activation sites on the LA remained consistent between monoatrial and biatrial configurations. A point was classified as inducing if reentry was initiated and maintained for at least 1 s. The vulnerability ratio was defined as the number of inducing points divided by the number of stimulation points. The mean tachycardia cycle length (TCL) of the induced reentries was further assessed at the stimulation site.

### 2.5. Statistical Analysis

Data are reported as mean *±* SD. To evaluate statistical significance between the sample means, we conducted a two-sampled t-test. A *p*-value *<* 0.05 was considered statistically significant.

## 3. Results

The 8 biatrial anatomical models and the number of stimulation points in each chamber are shown in Figure 2. The mean fibrotic extent in the LA was 12.20% *±* 0.68 and 44.01% *±* 2.6, for the M and S states, respectively. For the RA, the fibrosis extent was 5.31% *±* 0.13 and 12.96% *±* 0.68 for M and S states, respectively. The amount of fibrosis for each subject in each stage is shown in Table S2 in the supplementary material.

**Figure 2:**
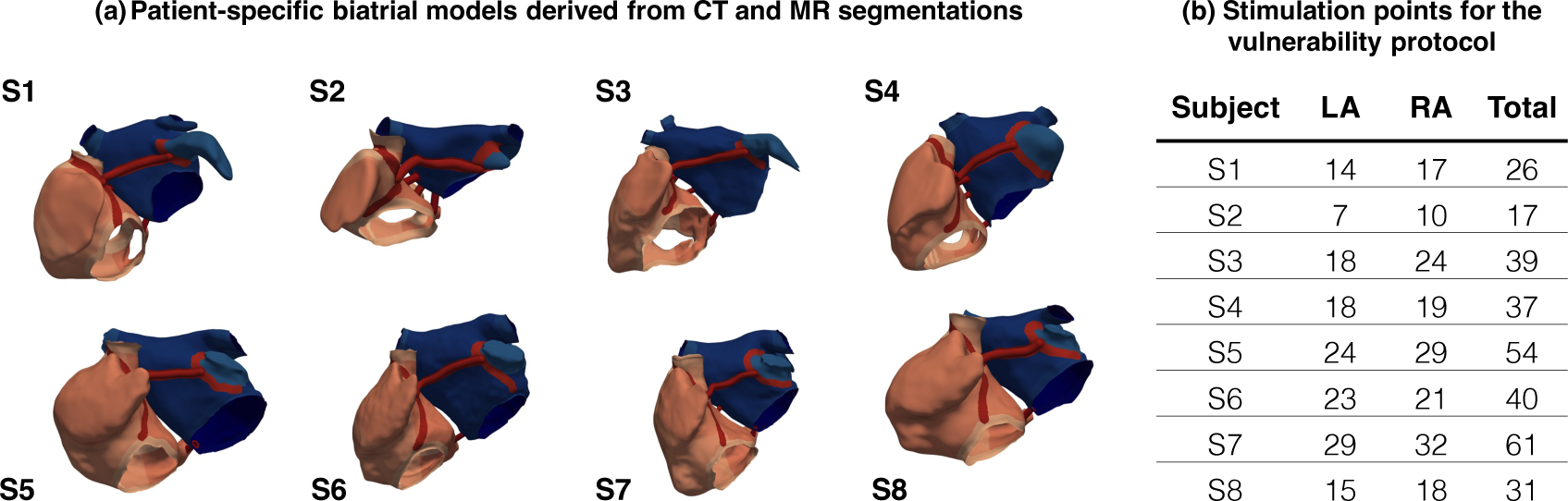
Personalized models and the total number of stimulation points used for the assessment of arrhythmia vulnerability. (CT: computed tomography, MR: magnetic resonance).

### 3.1. Monoatrial configuration

We ran 444 monoatrial simulations, from a total of 148 stimulation points x 3 remodeling states, to assess arrhythmia vulnerability in the 8 LA models. The number of inducing points and the vulnerability ratio *V*_LA_ for each subject are shown in Figure 3a. A total of 79 reentry episodes were induced, of which 32 episodes were in the M state and 47 in the S state. No reentries were induced in the H state. The vulnerability ratio *V*_LA_ among all subjects in the M and S states was 0.19 *±* 0.13 and 0.31 *±* 0.14, respectively. There was a 20.0% increase in the TCL between states M and S (186.94 *±* 13.3 vs. 224.32 *±* 27.6 ms), as illustrated in Figure 4a.

**Figure 3:**
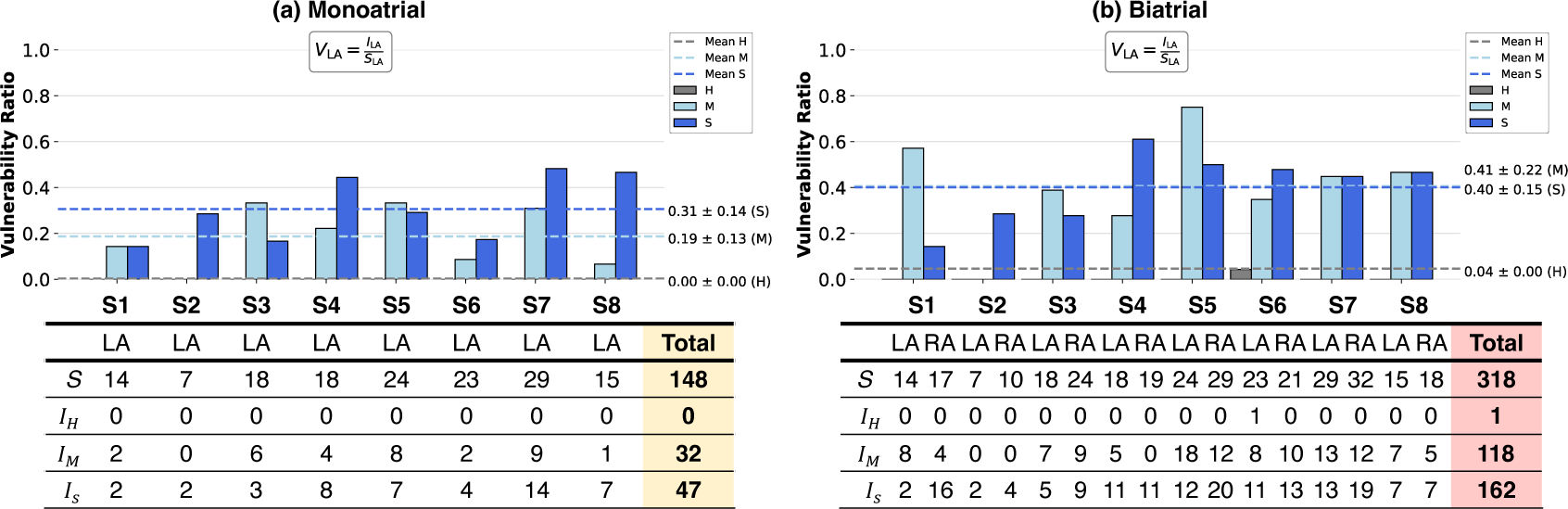
Vulnerability of the left atrium (LA) in monoatrial (a) and biatrial (b) configurations with respect to each remodeling scenario. H: healthy, M: mild, S: severe. *I_LA_* Inducing points in the LA, *S_LA_* Inducing points in the LA. The dashed lines represent the mean vulnerability ratio for each remodeling level.

**Figure 4:**
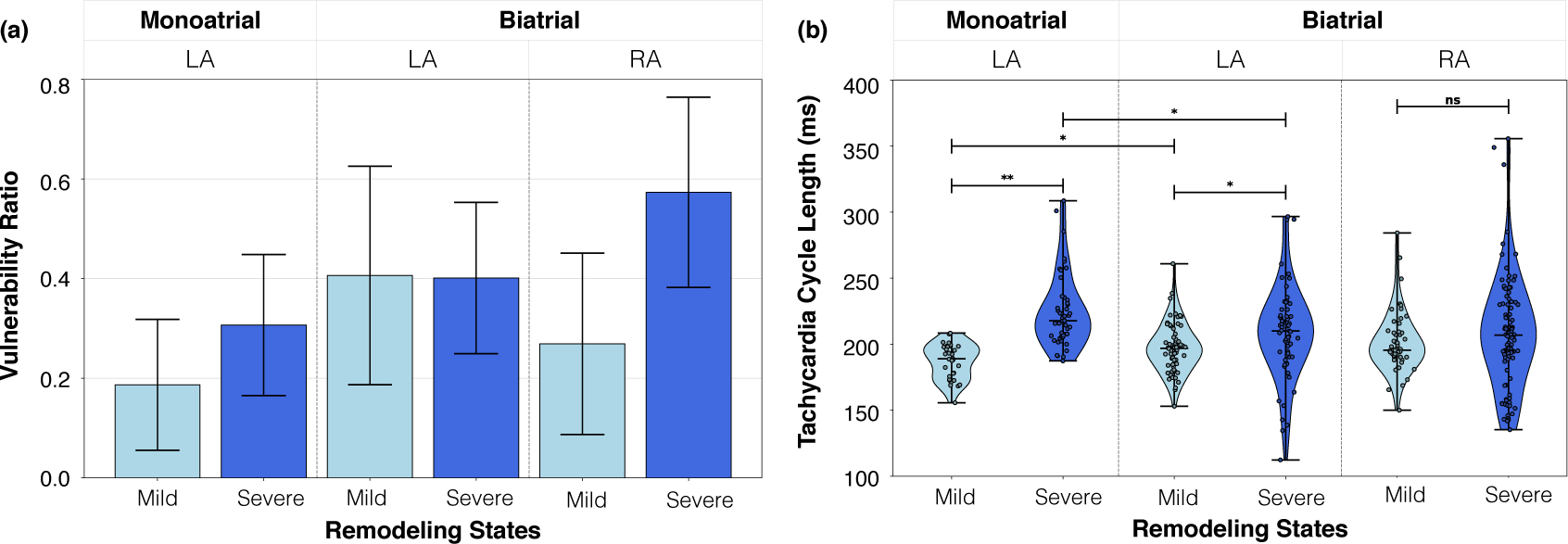
Impact of the right atrium (RA) on arrhythmia vulnerability ratio (a) and tachycardia cycle length (TCL) (b). Left atrium (LA) monoatrial refers to the LA in monoatrial configuration. Each bar corresponds to the vulnerability ratio, calculated as the number of induced points relative to the total points in each chamber across all 8 subject models. Each violin plot represents the probability density of TCL measurements. Medians are marked with horizontal lines inside the violins. Scatter points on top represent each reentry measurement. * p-value *<*0.05 is considered statistically significant ** p-value *<*0.01 (ns: not statistically significant).

The number of inducing points was higher in state S than M for 5 subjects (S2, S4, S6, S7, and S8), while for 2 of them (S3 and S5), the number of inducing points was higher in the M state. One subject (S1) exhibited no change in vulnerability ratio when increasing substrate extent from M to S. In state M, subjects S3 and S5 had the highest *V*_LA_ = 0.33, while in state S, S7 had the highest *V*_LA_ = 0.48. Increased remodeling from M to S revealed 4.3 *±* 2.9 new inducing points in the LA per patient, as shown in Figure 5a. The points became inducing when going from M to S due to rotational activity near the fibrotic regions. Deceleration of the wavefront and a shortened action potential in S enabled propagation within the fibrotic region. In contrast, in M, the faster wavefront encountered refractory tissue and failed to activate the surrounding tissue.

**Figure 5:**
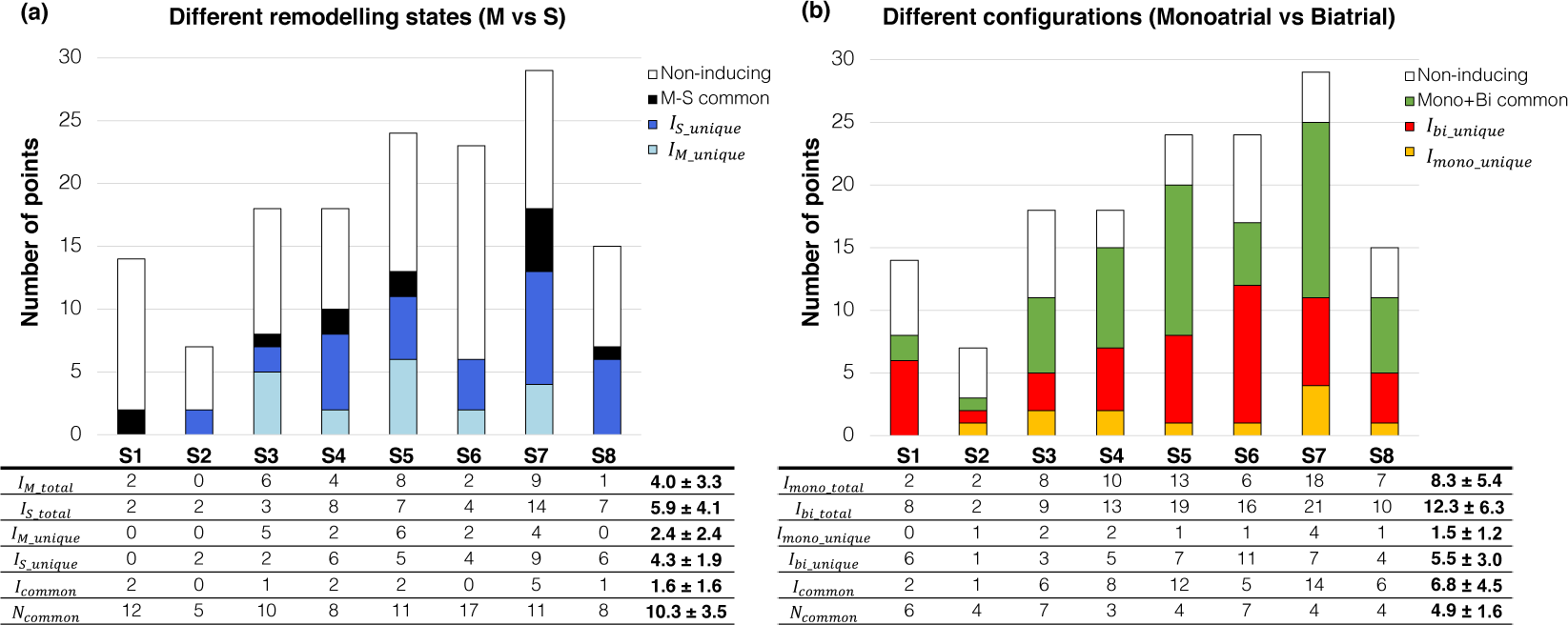
Proportion of inducing (I) and non-inducing (N) points in the left atrium (LA) shows higher inducibility due to increased remodeling in the monoatrial (mono) configuration (a) and higher inducibility due to the incorporation of the right atrium in biatrial (bi) configuration (b). Unique refers to points that exclusively induce in a specific setup. (M: mild, S: severe).

### 3.2. Biatrial configuration

A total of 954 biatrial simulations, from a total of 318 stimulation points x 3 remodeling states, were performed to evaluate vulnerability in the 8 biatrial models. The number of inducing points in both chambers are shown in Figure 3b and the vulnerability ratio in the biatrial scenario for each chamber is shown in Figure 4a. A total of 281 reentry episodes were induced in the biatrial configuration, of which 130 were induced by pacing from the LA and 151 by pacing from the RA. In the H state, only one reentry was induced by pacing from the anterior wall of the LA in the proximity of the mitral valve in S6.

The vulnerability ratio of the RA (*V*_RA_) is shown in Figure 4a. There was a 111.1% increase between states M and S in *V*_RA_ (0.27 *±* 0.18 vs. 0.57 *±* 0.19). The mean TCL of the RA-induced reentries for the M and S state was 201.33 *±* 23.0 ms and 207.87 *±* 41.6 ms (*p* = 0.295), as shown in Figure 4b. The vulnerability ratio of LA-induced reentries between the M and S states showed minimal changes (0.41 *±* 0.22 vs. 0.40 *±* 0.15). The mean TCL of LA-induced reentries in the biatrial configuration showed a significant difference between the M and S states (197.24 *±* 18.3 ms vs. 208.24 *±* 34.8 ms, *p* = 0.026).

### 3.3. Right atrium

To assess the role of the RA on LA inducibility in more detail, we additionally compared the arrhythmia vulnerability ratio and the TCL of the LA in monoatrial vs. biatrial configurations. The incorporation of the RA led to an increase in the mean LA vulnerability from 0.19 *±* 0.13 to 0.41 *±* 0.22 in state M and from 0.31 *±* 0.14 to 0.40 *±* 0.15 in state S, as shown in Figure 4a. Additionally, including the RA led to changes in the TCL of the LA-induced reentries in the M scenario from 186.94 *±* 13.3 ms to 197.24 *±* 18.3 ms (*p* = 0.006) and in the scenario S from 224.32 *±* 27.6 ms to 208.24 *±* 34.8 ms (*p* = 0.010).

Furthermore, we assessed changes in the inducibility of the LA by comparing points within the LA initiating reentry with and without the RA, as shown in Figure 5b. The inclusion of the RA resulted in an elevated LA inducibility, uncovering 5.5 *±* 3.0 inducing points in the LA biatrial scenarios that did not induce in the monoatrial configuration, as shown in Figure 6. The IAC at the CS contributed to the increased reentry inducibility, as shown in Figure 7.

**Figure 6:**
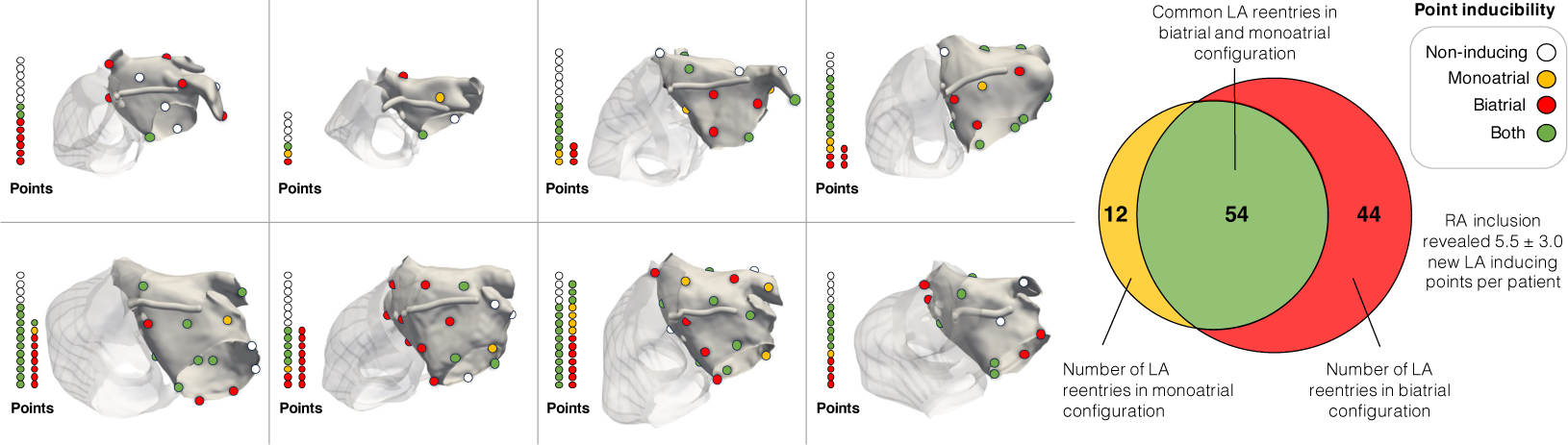
Increased inducibility in the left atrium (LA) due to incorporation of the right atrium (RA). The meshes show the stimulation points in the LA inducing reentry in monoatrial (yellow), biatrial (red), both (green) configurations, or non-inducing (white). The columns of points illustrate the type of inducibility at each stimulation point for each subject. The Venn diagram (right) depicts monoatrial and biatrial reentry distribution among all subjects. The RA appears attenuated.

**Figure 7:**
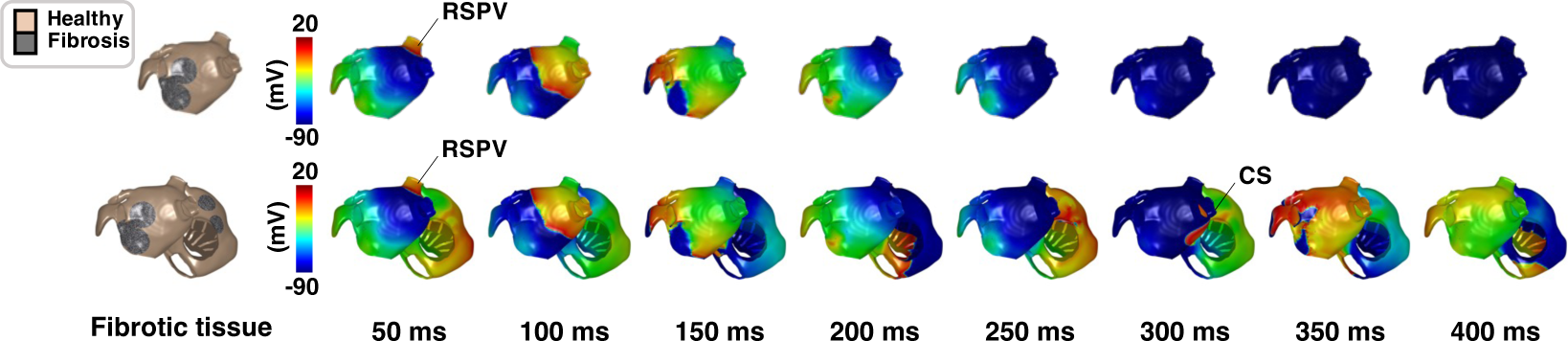
Reentry induction in biatrial configuration aided by inter-atrial connections. View of the posterior wall of the left atrium (LA). Stimulation point at the right superior pulmonary vein (RSPV) of the LA, the reentrant pathway involved the inter-atrial connection via the coronary sinus (CS) and fosa ovalis (not visible), and was therefore not supported by the monoatrial setup.

## 4. Discussion

In this study, we evaluated 2 different chamber scenarios: monoatrial and biatrial; and 3 different remodeling states: H, M, and S; in 8 different patient-specific anatomical models resulting in 48 arrhythmia vulnerability assessments. The main focus was to assess the role of the RA for arrhythmia vulnerability in a cohort of patient-specific anatomical models. Our study provides evidence for the importance of the RA for the maintenance and induction of arrhythmia and a reduction of the effect of substrate remodeling in biatrial settings.

### 4.1. Impact of the right atrium

The notion that the RA could play a role in AF is not a novel concept, as indicated by Nitta et al. [45]. However, the existing literature often neglects this potential role and provides limited evidence regarding the extent to which the RA contributes to the initiation and maintenance of AF. Scrutinizing the latest guidelines for AF treatment reveals the absence of the term “right atrium” [46, 47]. This highlights a scarcity of comprehensive studies investigating the role of the RA in the context of AF prevention and treatment.

To study the role of the RA as a potential initiator of or contributor to AF, we evaluated the RA vulnerability ratio. Among all investigated configurations, the RA was the chamber with the highest vulnerability in the S state. A possible explanation could be the larger RA size and the increased electrophysiological heterogeneity due to the presence of the PM, CT, and TV. Despite the lower fibrotic extent in the RA compared to the LA, the RA was more vulnerable to developing reentry upon stimulation than the LA.

In our simulations, we identified additional inducing points in the LA biatrial configuration which did not induce reentry in the LA-only model,as shown in Figure 7. One hypothesis suggests that the IACs play an important role in the maintenance of arrhythmia as mentioned by Roney et al. [43]. In terms of arrhythmia mechanisms, for reentry to occur, there must be an excitable gap, meaning that the wavelength must be shorter than the reentrant circuit length [48]. The inclusion of the RA increased the likelihood of new reentrant circuits influenced not only by larger atrial size but also CV and effective refractory period [49].

Previous computational model studies have established that the dynamics of reentrant drivers are influenced by the extent and distribution of the fibrotic substrate, in the RA [50] and LA [51]. Moreover, investigations by Boyle et al. [19] and Zahid et al. [50] have identified reentrant drivers in the RA through the utilization of biatrial models. In our simulations we also observed simultaneous interactions of multiple reentries (functional and anatomical) in our biatrial simulations, such as rotational activity around the tricuspid and mitral valves, unidirectional blocks in the BB region, reentrant pathways aided by IACs, and rotors associated with the fibrotic substrate. In summary, we propose that the increased inducibility in the LA biatrial model, i.e. additional reentrant drivers, resulted from the interplay between fibrosis characteristics and novel circuit paths, as shown in Figure 8.

**Figure 8:**
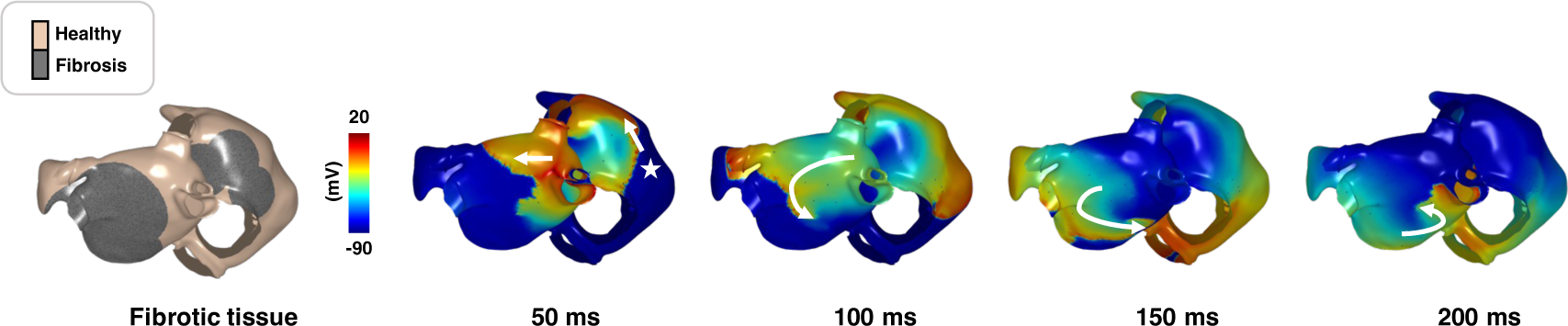
Example of reentry induction from stimulation point in the right atrium (RA) in S state. The distribution of fibrosis is shown on the left. The inducing point (star) is located in the RA near the inferior vena cava. The reentry is anchored at the inferior wall of the left atrium and the wave propagation slows down at the border of the fibrotic region.

An additional finding concerns changes in the LA vulnerability and the TCL distribution. Without the RA, the vulnerability of the LA was markedly higher in the S than in the M state. Once the RA was incorporated, this difference of LA vulnerability was markedly reduced. In terms of reentry dynamics, adding the RA increased the mean TCL of LA reentries in the M state by 5.21% (186.94 *±* 13.3 vs. 197.24 *±* 18.3), e.g. causing slower reentries, and on the contrary, the RA led to faster LA reentries in the S state (224.32 *±* 27.6 vs. 208.24 *±* 34.8 ms). The observed changes suggest a state-dependent influence of the RA on reentry dynamics in the LA. These results might be due to the TCL of the LA in the biatrial configuration being similar to the TCL of the RA. This could explain the state-dependent effect of the RA as the detected increase in the TCL in the LA might be due to additional reentrant activity promoted by the RA substrate, rather than the LA. Also, faster reentries in the S state in the biatrial configuration could be attributed to the lesser fibrotic burden in the RA, therefore allowing faster propagation.

One of the main implications of these findings lies in the development of tools informing ablation therapy using computer models. It can be expected that the arrhythmia vulnerability ratio calculated based on LA-only models changes once the RA is incorporated. This is especially relevant as successful virtual ablation therapies for AF are based on non-inducibility criteria. Therefore, performing biatrial simulations appears advisable. In summary, the incorporation of the RA increased the vulnerability of the LA and the increase in substrate extent had a lesser effect on the vulnerability of the LA.

### 4.2. Arrhythmia vulnerability in different remodeling states

The majority of subject models exhibited higher vulnerability in the S state. Yet for some, the vulnerability ratio was higher in the M state. To understand this behavior, we analyzed activation patterns of reentries induced only in the M state. In the M state, the fibrotic substrate impeded wavefront propagation, causing unidirectional blocks and anchoring reentries. Conversely, in the S state, increased fibrosis led to a slower wavefront progression, facilitating tissue recovery and promoting regular activation. For the other cases where the S state had a higher vulnerability, the faster wavefront in M encountered refractory tissue and failed to activate the surrounding tissue. While in S, deceleration of the wavefront and a shortened action potential enabled propagation within the fibrotic region. The overall outcome was a combination of both effects.

### 4.3. Limitations

To our knowledge, this study represents the first dedicated examination of the role of the RA in arrhythmia vulnerability in patient-specific computer models, however the limited sample size may impact the generalization of our findings. The IACs were modeled as structures connecting the LA and RA implemented using rule-based definitions [25]. It is possible that different IAC configurations, including different numbers of functional IACs and different locations, might affect the maintenance of reentrant pathways. All virtual patient models had a similar fibrosis pattern based on circular patches. Future studies could include more complex patterns of fibrosis [52] to study their impact on the vulnerability of biatrial models.

## 5. Conclusions

LA reentry vulnerability in a biatrial model is higher than in a monoatrial model. Incorporating the RA in patient-specific computational models unmasked potential inducing points in the LA. The RA had a substrate-dependent effect on reentry dynamics and affected the TCL of the LA-induced reentries. As virtual ablation strategies for AF are based on non-inducibility criteria, performing biatrial simulations is advisable. Our study highlights the importance of the RA for the maintenance and induction of arrhythmia in patient-specific computational models.

## Funding

This project has received funding from the European Union’s Horizon 2020 research and innovation programme under the Marie Sklodowska-Curie grant agreement No 860974. This work was supported by the Leibniz ScienceCampus “Digital Transformation of Research” with funds from the programme “Strategic Networking in the Leibniz Association”. The authors acknowledge support by the state of Baden-Württemberg through bwHPC.

## Data Availability

A carputils bundle containing
the openCARP experiment is publicly available https://dx.doi.org/10.35097/1830

## Notes

### Competing Interest Statement

The authors have declared no competing interest.

### Author Declarations

Subjects provided written informed consent and the study protocol was reviewed and approved by the ethical committee of Guy's Hospital, London, UK, and University Hospital Heidelberg, Heidelberg, Germany

