## Supplementary_Material for "The Right Atrium Affects in silico Arrhythmia Vulnerability in Both Atria"

| | S1 | S2 | S3 | S4 | S5 | S6 | S7 | S8 |
|---|---|---|---|---|---|---|---|---|
| Sex | F | F | M | M | M | M | M | M |
| Diagnosis | Ctl | LQT2 | Ctl | LQT1 | Ctl | Ctl | AF | Ctl |
| HR (1/min) | 81 | 76 | 69 | 62 | 70 | 53 | 62 | 86 |
| PWd (ms) | 95 | 95 | 107 | 91 | 103 | 97 | 176 | 99 |
| RA blood volume (ml) | 98 | 52 | 117 | 88 | 132 | 99 | 155 | 72 |
| LA blood volume (ml) | 55 | 27 | 63 | 79 | 81 | 87 | 136 | 53 |
| RA myocardium ($mm^3$) | 26 | 12 | 27 | 38 | 52 | 21 | 38 | 36 |
| LA myocardium ($mm^3$) | 19 | 10 | 25 | 32 | 26 | 19 | 34 | 29 |

**Table S1:** Clinical characteristics of subjects for the generation of biatrial models. (F: Female, M: Male, HR: Heart rate, PWd: P-wave duration, RA: right atrium, LA: left atrium, Ctl: control; LQT: long-QT syndrome, AF: atrial fibrillation).

| | S1 | | S2 | | S3 | | S4 | | S5 | | S6 | | S7 | | S8 | |
|---|---|---|---|---|---|---|---|---|---|---|---|---|---|---|---|---|
| Utah stage | 2 | 4 | 2 | 4 | 2 | 4 | 2 | 4 | 2 | 4 | 2 | 4 | 2 | 4 | 2 | 4 |
| LA (%) | 10.9 | 36.4 | 12.1 | 42.3 | 12.7 | 35.4 | 12.7 | 38.3 | 13.0 | 34.8 | 11.7 | 40.3 | 13.1 | 41.9 | 11.4 | 41.7 |
| RA(%) | 5.4 | 12.8 | 5.3 | 13.3 | 5.3 | 13.4 | 5.9 | 14.1 | 5.36 | 11.7 | 5.3 | 13.4 | 5.4 | 13.1 | 5.4 | 11.9 |

**Table S2:** Percentage of fibrosis included for each remodeling level. The ranges of the percentage of fibrosis in each chamber are reported in [37]. Utah stage 2 corresponds to the M state and Utah stage 4 corresponds to the S state. ( RA: right atrium, LA: left atrium).

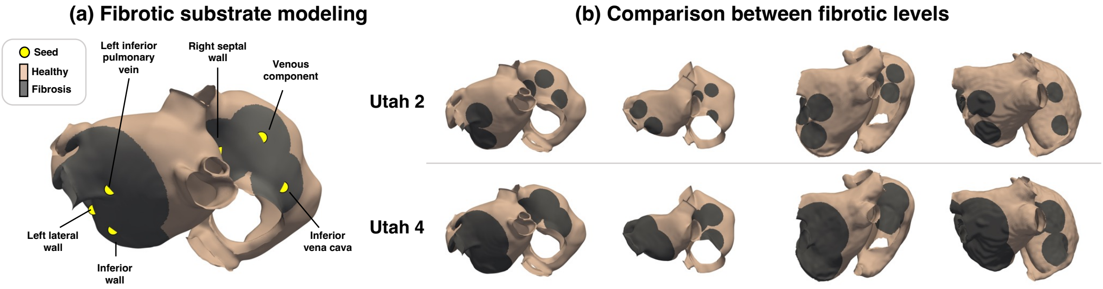


**Figure S1:** Fibrotic substrate modeling. (a) A total of 6 seeds were placed in each biatrial geometry to generate fibrosis distributions corresponding to the clinical Utah stages [41]. (b) Fibrosis distributions in 4 subjects in state M (top, Utah 2) and S (bottom, Utah 4).

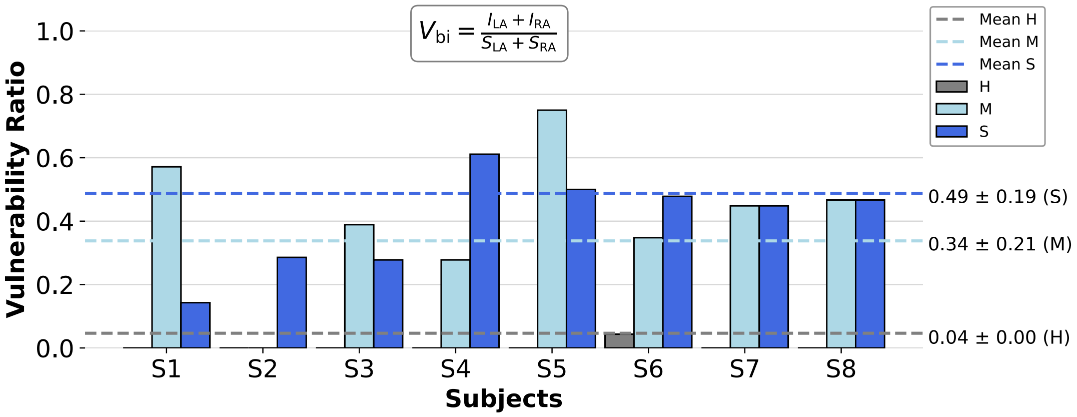


**Figure S2:** Vulnerability of the right atrium (RA) and left atrium (LA) in biatrial configuration. Number of inducing points in each chamber for each subject in biatrial configuration with respect to each remodeling scenario. H: healthy, M: mild, S: severe. $V_{\text{bi}}$: biatrial vulnerability , $I_{\text{RA}}$: Inducing points in the RA, $S_{\text{RA}}$: stimulation points in the RA, $I_{\text{LA}}$: Inducing points in the LA, $S_{\text{LA}}$: stimulation points in the LA.
